# Ordinal Probit Modeling of Loneliness, Relationship Difficulty, Worthlessness, and Depression in Midlife: An Ordinal Association Analysis of Psychosocial and Demographic Predictors

**DOI:** 10.64898/2026.11.25.26368969

**Authors:** Grace Katuri, Atchyuth Kumar Pairdha

## Abstract

Midlife is a time of significant social, economic, and health change, and the covariation between demographic and psychosocial characteristics and ill-being has both preventional and organisational implications. This is an attempt to produce a single ordinal variable summarising information about mental health-ill-being, and its dependence on demographic predictors. We measure mental ill-being through four midlife-related outcomes—loneliness frequency (RA1SA20G), ability to sustain close relationships (RA1SF1J), worthlessness frequency (RA1SA20F), and degree of depressive symptomatology (RA1PDEPRE)—and explore a fifth, anxiety (RA1PANXIE) at the descriptive, but not inferential, level. The predictors are age and sex, pain of financial difficulty in meeting the monthly bills, purposefulness/agentic engagement, self-efficacy, sleep difficulty, contact with social media, meditation/relaxation use, and leisure vigorous activity. After removing missing or refused values (‘listwise deletion’), 2,423 respondents (52.9% female; age range 23–76, mean 51.84, SD 14.20) from the initial 3,577 sample extract remain; we give univariate, one-variable summaries, merge sparse categories of the predictor variables through a limited number of theoretically motivated recodes (reversing the disagreement coding, amalgamating the five frequency levels, trichotomising the depression variable (none, average, or severe), and binning age into quartiles), and establish the strength of association of predictor pairs with ordinal correlation coefficients (Kendall’s tau-b, Stuart’s tau-c, and rank-biserial correlation for sex). We fit maximum-likelihood ordinal probit models both as models with a sole predictor variable (each in turn) and as a conditional (multi-predictor) model with four predictors (bins of age, bill difficulty, purposefulness/agency, and sleep difficulty) as a function of cross-validated predictive accuracy estimates from stratified five-fold cross-validation. We find high cross-validated predictive accuracy for depression (0.864), and worthlessness (0.808), and lower but still modest predictive accuracy for loneliness (0.653–0.655) and the ability to sustain close relationships (0.291–0.304); single-predictor accuracy estimates were not improved substantially in the combined models for loneliness (0.653–0.654) and ability to maintain close relations (0.291–0.304). The plots of the cumulative association functions show perfect monotonic relationships— more financial difficulty and disrupted sleep lead to greater probability of an adverse outcome—while the ordinal association plots show that in general the predictors are not qualitatively different from one another. The partial derivatives of the predicted cumulative functions indicate that the covariation between these predictor variables and mental ill-being operates in common along monotonic, ordinal–ordinal continua. We discuss how our decisions about recode levels, the interpretation of the ordinal association coefficients, potential limitations of the models (such as bias introduced through codings and the treatment of ordinal scales as equidistant, as well as the failure to treat sex as a nominal variable) and how future predictions could benefit from the inclusion of sample weights, ordinal-loss functions for model assessment, ordinal proportional-odds diagnostics, and model-based causal pathways from longitudinal waves of the MIDUS study.

## 1. Introduction and Midlife Mental Health Context

Adulthood between emerging workforce power and late-life retirement involves changing occupational, caregiving, and house-hold spending burdens and changing social connections. In this stage of life, mental health experiences tend to be multidimensional rather than uni-dimensional symptom counts (1). Examination of which demographic factors and psychosocial resources align with ordered affective levels enables an understanding of routes to risk across midlife samples (2).

These relationships can be modeled in large nationally representative survey programs using structured survey questions. A nationally representative survey platform that includes measured variables on psychological, interpersonal, sleep and economic stress, as well as measures of psychological functioning is the Midlife in the United States (MIDUS) study(3). With access to Project 1 data, one can assess the marginal severity distributions of survey responses as well as the multivariable relationships between ordered survey responses.

Three broader issues are associated with the use of indicators-discrete “rough metrics”-from a database for the purpose of building models and inferences. First, this paper draws on a curated basket of survey indicators directly availiable from the MIDUS Project 1 and examining these four major outcomes during their inferential and predictive modeling: (1) frequency of feeling lonely (`RA1SA20G`); (2) difficulty and frustration with close relationships (`RA1SF1J`); (3) frequency of feeling worth-less (`RA1SA20F`); and (4) an affect-plus-anhedonia depression symptom score (`RA1PDEPRE`). An anxiety score (`RA1PANXIE`) is examined during profile displays but not during the post-recode modeling targets.

Candidate predictors include demographic background, financial stress, perceived agency, sleep disturbance, digital social contact, engagement in stress-management technique, and physical activity. The feature set specifically comprises age of respondent (`RA1PRAGE`) and respondent sex (`RA1PRSEX`), difficulty in paying monthly bills (`RA1SH9`), purpose/agency (`RA1SF100`), self-efficacy (`RA1SF4F`), difficulty falling asleep (`RA1SA57A`), social media contact with friends (`RA1SK6`) and non-coresident family members (`RA1SK2`), engaging in relaxation or meditation (`RA1SA52N`), and vigorous summer leisure activity (`RA1SA26E`). Self-efficacy and social support have been jointly studied for their effects on loneliness and depression in older adults with multimorbidity (4).

The empirical analysis is framed by three questions. How do the un- and recoded categories of these demographic, psychosocial and affective measures behave across the analytic sample? What bivariate rank association patterns link predictors and out-comes when score tests are performed with concordance-based indices and rank-biserial correlation? How well do single- and four-predictor ordinal probit models classify held-out observations under stratified five-fold cross-validation?

Fig. 1 brings together the analysis pipeline, which progresses from data collection to cross-validated ordinal probit modeling. Instead of collapsing the ordered self-report ratings into an un-constrained continuous scale, the workflow retains the ordering of categories while accounting for idiosyncratic tail cell sparsity using transparent recoding rules. It combines non-parametric rank summaries with parametric latent-threshold modeling to examine results at the margins as well as for outcome classifications. An analogous interpretable analytic pipeline was described for lifestyle-based diabetes screening (5).

**Figure 1:**
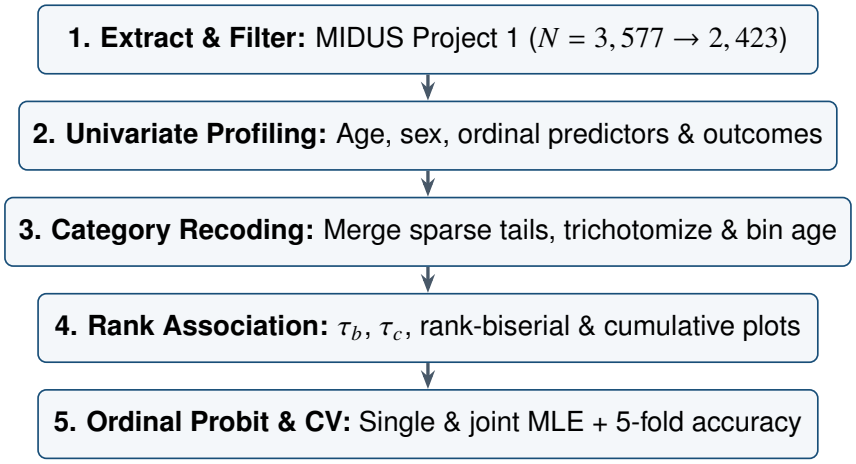
End-to-end analytical workflow applied to the MIDUS Project 1 dataset.

The remaining sections keep the paper structured. Section II explains how the sample is assembled, how variables are constructed and categorization is recoded. Section III specifies the non-parametric association measures, ordinal probit model specification, and cross-validation procedure. Section IV displays sample means, post-recode distributions, pairwise associations, cumulative thresholds and accuracy scores. Section V discusses how substantive and methodological trade-offs lead to recommendations, and Section VI concludes and suggests extensions.

### 2. Data Architecture, Sample Construction, and Category Recoding

The empirical data table is from a tabular view of MIDUS Project 1 (`midus_raw.csv`), with 3, 577 starting records from adult study participants. Administrative variables (`SAMPLMAJ` and `MRID`) are removed in preprocessing, leaving participant key (`MIDUSID`), demographic, predictor, and outcome columns. Complete-case filtering deletes any record with a missing or refused value on an analytic variable, resulting in a modeling data table (`dataset.csv`) with *N* = 2, 423 respondents and 15 ordered columns.

Table 1 consolidates sample attrition and baseline demographic distributions. Women account for 52.9% (*n* = 1, 281) of the analytic cohort and men account for 47.1% (*n* = 1, 142). Chronological age spans 23 to 76 years with a mean of 51.84 (SD = 14.20), a median of 52.0, a mode of 43, and an interquartile range of 24.0 years (*Q*_1_ = 40.0, *Q*_3_ = 64.0). Slight negative skewness (−0.11) and platykurtic kurtosis (−1.20) reflect a broad spread across young-adult, core midlife, and older-adult age bands.

**Table 1:**
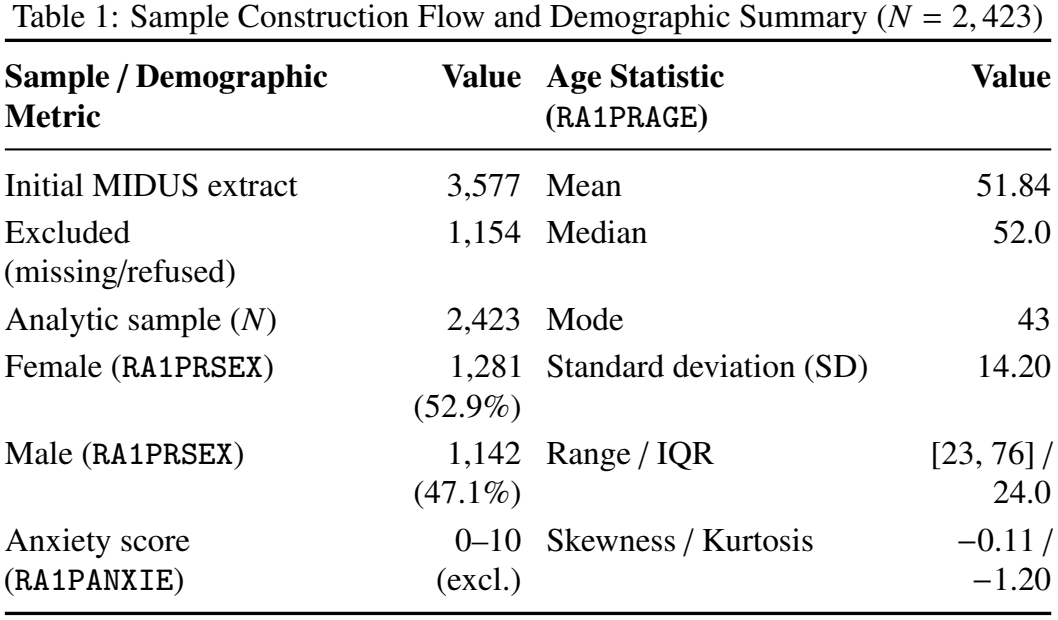
Sample Construction Flow and Demographic Summary (*N* = 2, 423)

| **Sample / Demographic Metric** | **Value** | **Age Statistic (RA1PRAGE)** | **Value** |
|---|---|---|---|
| Initial MIDUS extract | 3,577 | Mean | 51.84 |
| Excluded (missing/refused) | 1,154 | Median | 52.0 |
| Analytic sample (*N*) | 2,423 | Mode | 43 |
| Female (RA1PRSEX) | 1,281 (52.9%) | Standard deviation (SD) | 14.20 |
| Male (RA1PRSEX) | 1,142 (47.1%) | Range / IQR | [23, 76] / 24.0 |
| Anxiety score (RA1PANXIE) | 0–10 (excl.) | Skewness / Kurtosis | −0.11 / −1.20 |

Raw survey items capture distinct domains across multi-point ordered scales. Financial strain (`RA1SH9`) rates difficulty paying monthly bills across four ordered levels, while sleep onset trouble (`RA1SA57A`) and relaxation or meditation use (`RA1SA52N`) use five-level frequency scales. Purpose/agency (`RA1SF100`) and self-efficacy (`RA1SF4F`) measure agreement on seven-point scales, social media contact with friends (`RA1SK6`) and family (`RA1SK2`) span eight frequency levels, and vigorous summer leisure activity (`RA1SA26E`) spans six frequency levels. Multi-level ordinal instruments of this kind are also common in clinical outcome research (6).

On the outcome side, loneliness (`RA1SA20G`) and worthlessness (`RA1SA20F`) ask how often respondents experienced each feeling during the past 30 days, whereas relationship difficulty (`RA1SF1J`) asks how strongly respondents agree that maintaining close relationships has been difficult and frustrating. The depression composite (`RA1PDEPRE`, ranging from 0 to 7) aggregates depressed affect and anhedonia, while the anxiety score (`RA1PANXIE`, ranging from 0 to 10) is kept for descriptive inspection rather than entered into the post-recode probit models. The relationship between such measures and access to care has been considered in prior youth mental health work (7).

Table 2 specifies the category transformations applied before bivariate association and regression modeling. Sparse lower-end disagreement categories on self-efficacy (`RA1SF4F`) and purpose/agency (`RA1SF100`) are combined into a single disagreement tier to stabilize cell counts. High-frequency upper categories are merged for relaxation/meditation (`RA1SA52N`), loneliness (`RA1SA20G`), and worthlessness (`RA1SA20F`) where extreme cells are comparatively small.

**Table 2:**
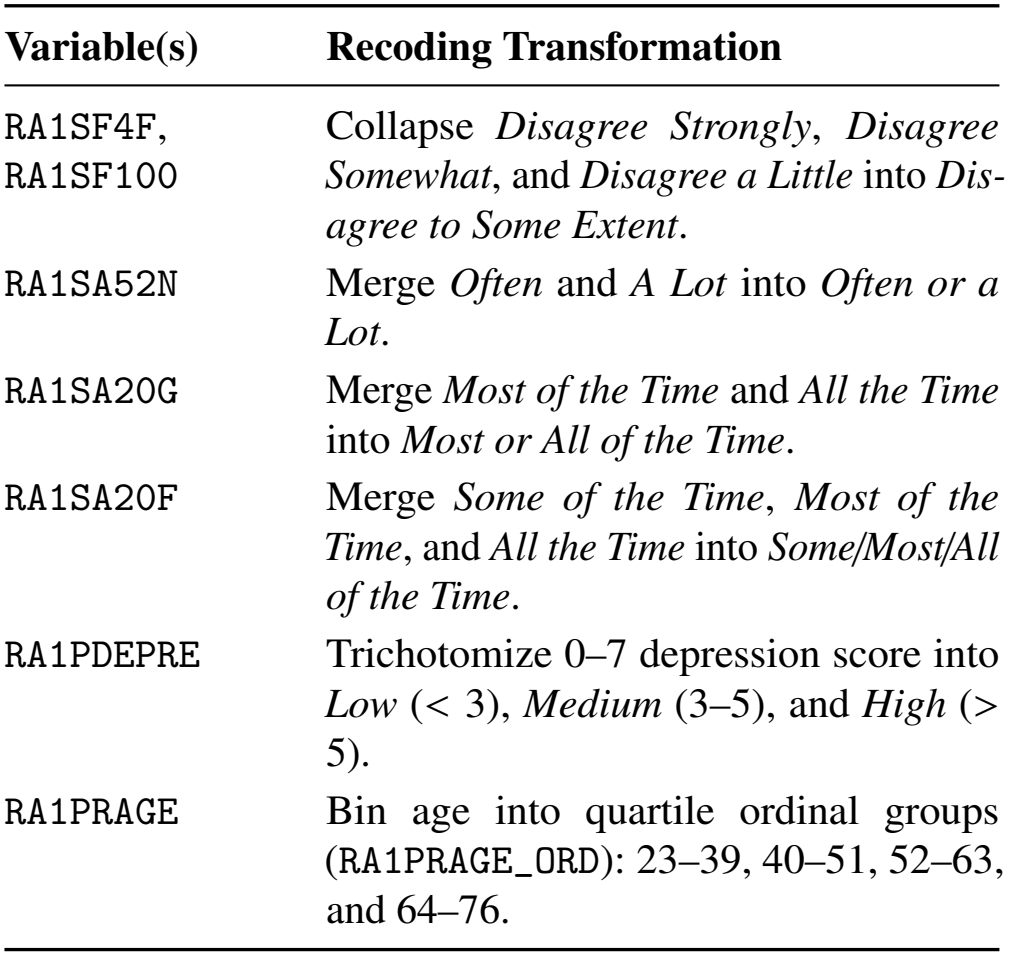
Category Recoding Rules Applied Prior to Association and Probit Modeling.

| **Variable(s)** | **Recoding Transformation** |
|---|---|
| RA1SF4F, RA1SF100 | Collapse *Disagree Strongly*, *Disagree Somewhat*, and *Disagree a Little* into *Dis-agree to Some Extent*. |
| RA1SA52N | Merge *Often* and *A Lot* into *Often or a Lot*. |
| RA1SA20G | Merge *Most of the Time* and *All the Time* into *Most or All of the Time*. |
| RA1SA20F | Merge *Some of the Time*, *Most of the Time*, and *All the Time* into *Some/Most/All of the Time*. |
| RA1PDEPRE | Trichotomize 0–7 depression score into *Low* (< 3), *Medium* (3–5), and *High* (> 5). |
| RA1PRAGE | Bin age into quartile ordinal groups (RA1PRAGE_ORD): 23–39, 40–51, 52–63, and 64–76. |

Continuous or count-like features are likewise mapped onto compact ordinal scales for unified modeling. The numeric depression score (`RA1PDEPRE`) is grouped into three severity tiers—low (< 3), medium (3–5), and high (> 5)—creating a clear ordinal target structure. Continuous age (`RA1PRAGE`) is partitioned into four quartile-aligned intervals (`RA1PRAGE_ORD`: 23–39, 40–51, 52–63, and 64–76) so that age can enter both rank correlation tables and ordinal probit specifications alongside discrete predictors.

## 3. Non-Parametric Association, Ordinal Probit Specification, and Evaluation

Once raw codes are mapped to labeled categories and filtered to complete cases (*N* = 2, 423), pairwise bivariate relationships are evaluated across sex, quartile-binned age, recoded predictors, and recoded outcomes. Rather than relying on Pearson productmoment correlations that impose linear interval spacing, the analysis pairs concordance-based rank correlations for ordered pairs with a rank-biserial measure for the dichotomous sex indicator. Statistical significance is flagged at the *p* < 0.05 threshold across heatmap cells. Rank and latent-variable approaches are also used in financial time-series research (8).

For two ordinal variables sharing the same number of categories, pairwise association is quantified using Kendall’s τ_*b*_, which adjusts for tied ranks on both marginals:

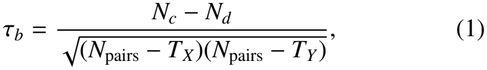

where *N*_*c*_ denotes concordant pairs, *N*_*d*_ denotes discordant pairs, 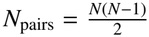 is the total number of distinct respondent pairs, and *T*_*X*_ and *T*_*Y*_ count pairs tied on the first and second variable, respectively.

When two ordinal variables differ in category counts and form a rectangular contingency table with min(*r, c*) dimensions, Stuart’s τ_*c*_ scales pair differences by the smaller table dimension:

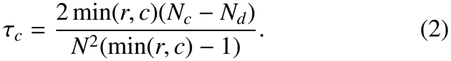

When one variable is dichotomous respondent sex (`RA1PRSEX`), association with an ordered variable is summarized via a rank-biserial correlation derived from the Mann–Whitney *U* statistic, providing a signed rank comparison between female and male subsamples bounded between −1 and +1.

To model conditional shifts across ordered outcome categories, we estimate ordinal probit regressions via maximum likelihood (`statsmodels.miscmodels.OrderedModel` with a probit link), illustrated in Fig. 2. For respondent *i*, an unob-served continuous propensity 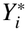 is linked to covariates *x*_*i*_ and standard normal disturbances:

**Figure 2:**
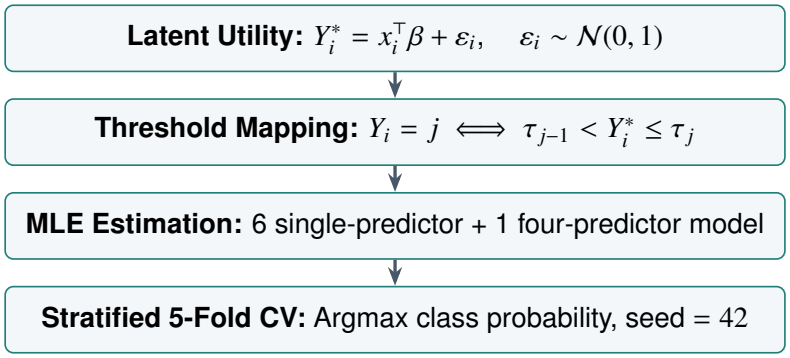
Latent-variable formulation, maximum likelihood estimation, and cross-validation design.

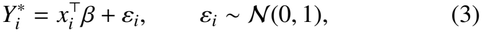

and the observed ordinal outcome *Y*_*i*_ *∈* {0, 1, … , *J*} falls into category *j* according to strictly increasing cutpoints τ_−1_ = −∞ < τ_0_ < τ_1_ < · · · < τ_*J*−1_ < τ_*J*_ = +∞:

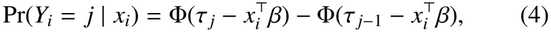

where Φ(·) denotes the standard normal cumulative distribution function.

Ordered predictors enter regression equations as integer codes preserving their natural rank progression, while respondent sex enters as a binary indicator with one reference category omitted. For each of the four primary outcomes (`RA1SA20G`, `RA1SF1J`, `RA1SA20F`, and `RA1PDEPRE`), we fit six single-predictor models—covering quartile-binned age, bill difficulty, purpose/agency, self-efficacy, sleep trouble, and sex— alongside one joint model containing four core predictors: binned age (`RA1PRAGE_ORD`), bill difficulty (`RA1SH9`), purpose/agency (`RA1SF100`), and sleep trouble (`RA1SA57A`).

Out-of-sample classification performance is evaluated using stratified five-fold cross-validation (*k* = 5, random seed 42) so every fold mirrors the outcome category proportions of the full sample. Within each split, parameters are estimated on the training folds and held-out observations are assigned the category maximizing predicted probability, 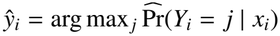, with performance averaged across folds. In addition, cumulative association plots display empirical exceedance proportions 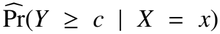 across predictor strata to inspect non-parametric monotonicity across outcome cutpoints. Cross-validated evaluation of this kind is standard practice for structured predictive models (9).

## 4. Empirical Results: Distributions, Associations, and Model Accuracy

Exploratory profiling of the 2, 423 complete-case respondents highlights pronounced concentration toward favorable or low-frequency states across most affective and behavioral items. In the raw data, loneliness (`RA1SA20G`) has a mode and median of 1 on its 1–5 scale with IQR = (1, 2), and worthlessness (`RA1SA20F`) similarly centers at 1 with IQR = (1, 1). Both the depression composite (`RA1PDEPRE`, range 0–7) and the anxiety score (`RA1PANXIE`, range 0–10) exhibit mode 0, median 0, and IQR = (0, 0).

Table 3 summarizes the post-recode structure of the primary outcomes alongside bill difficulty and quartile-binned age. After trichotomizing `RA1PDEPRE`, 86.4% (2, 093) of respondents fall into the low tier, 7.3% (177) into the medium tier, and 6.3% (153) into the high tier. Worthlessness (`RA1SA20F`) shows a similarly skewed distribution after merging upper frequency cells, with 80.8% (1, 957) reporting feeling worthless none of the time, 10.9% (264) a little of the time, and 8.3% (202) some, most, or all of the time.

**Table 3:** Post-Recode Distributions of Primary Outcomes and Key Predictors (*N* = 2, 423)

| **Variable** | **Category / Tier** | **Count** | **Share** |
|---|---|---|---|
| RA1SA20G<br>(Loneliness) | None of the Time | 1,583 | 65.3% |
|  | A Little of the Time | 462 | 19.1% |
|  | Some of the Time | 270 | 11.1% |
|  | Most or All of the Time | 108 | 4.5% |
| RA1SA20F<br>(Worthlessness) | None of the Time | 1,957 | 80.8% |
|  | A Little of the Time | 264 | 10.9% |
|  | Some/Most/All of the Time | 202 | 8.3% |
| RA1PDEPRE<br>(Depression) | Low (< 3) | 2,093 | 86.4% |
|  | Medium (3–5) | 177 | 7.3% |
|  | High (> 5) | 153 | 6.3% |
| RA1SH9<br>(Bill Difficulty) | Not at All Difficult | 742 | 30.6% |
|  | Not Very Difficult | 804 | 33.2% |
|  | Somewhat Difficult | 698 | 28.8% |
|  | Very Difficult | 179 | 7.4% |
| RA1PRAGE_ORD<br>(Age Bin) | 23–39 years | ∼606 | ∼25.0% |
|  | 40–51 years | ∼606 | ∼25.0% |
|  | 52–63 years | ∼606 | ∼25.0% |
|  | 64–76 years | ∼605 | ∼25.0% |

Other predictors display varied dispersion across raw scales before recoding. Difficulty paying monthly bills (`RA1SH9`) has mode and median 2 on a 1–4 scale (IQR = (1, 3)), while trouble falling asleep (`RA1SA57A`) has mode and median 2 on a 1–5 scale (IQR = (2, 3)). Purpose/agency (`RA1SF100`) and self-efficacy (`RA1SF4F`) skew strongly upward toward agreement (mode 7, median 6, IQR = (5, 7)), whereas relaxation/meditation (`RA1SA52N`) is heavily concentrated at its lowest category (mode and median 1, IQR = (1, 2)). Portable measurement tools have likewise been applied to diabetes-related analysis (10).

In the pairwise association matrix, positive coefficients indicate that higher predictor levels co-occur with greater outcome severity, while negative coefficients indicate inverse rank ordering. Stronger psychosocial agency and self-efficacy align negatively with loneliness, relationship difficulty, worthlessness, and depression, whereas greater financial strain and more frequent sleep trouble align positively with those adverse outcomes. Social media contact with friends and family correlates positively with each other, while demographic sex comparisons remain distinct from ordinal–ordinal rank correlations.

Cumulative association plots reinforce these rank patterns across outcome cutpoints without assuming parametric spacing, following the monotonic upward pattern illustrated schematically in Fig. 3. Across higher monthly bill difficulty (`RA1SH9`) and more frequent trouble falling asleep (`RA1SA57A`), exceedance probabilities Pr(*Y* ≥ *c*) rise steadily across both lower and higher severity thresholds for depression (`RA1PDEPRE`) and worthlessness (`RA1SA20F`). Conversely, older age quartiles (`RA1PRAGE_ORD`) show lower cumulative proportions of loneliness (`RA1SA20G`) relative to younger cohorts. Depression and loneliness have also been linked to glycemic outcomes among middle-aged and older adults with diabetes (11).

**Figure 3:**
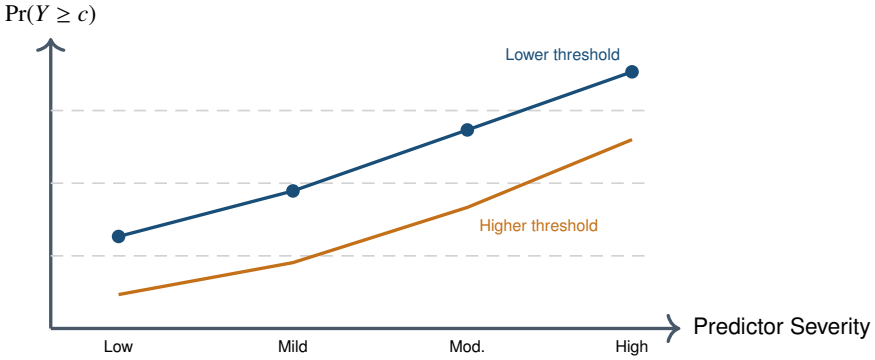
Schematic illustration of monotonic cumulative exceedance gradients across ordered outcome thresholds with increasing predictor severity.

Table 4 reports stratified five-fold cross-validated classification accuracies across all six single-predictor specifications. Accuracy is highest for depression (0.864) and worthlessness (0.808), intermediate for loneliness (0.653), and lowest for relationship difficulty (0.291–0.304). Notably, within each outcome column, accuracy remains nearly invariant across single predictors because majority-class assignment heavily influences hard classification when outcomes are skewed. Similar structural factors dominating outcome variation have been reported in organizational analyses (12).

**Table 4:** Mean Stratified 5-Fold Cross-Validation Accuracy of Single-Predictor Ordinal Probit Models.

| **Outcome** | **Age** | **Bill Diff.** | **Purpose** | **Self-Efficacy** | **Sleep** | **Sex** |
|---|---|---|---|---|---|---|
| Lonely | 0.653 | 0.653 | 0.653 | 0.653 | 0.653 | 0.653 |
| Rel. Diff. | 0.297 | 0.291 | 0.304 | 0.300 | 0.291 | 0.297 |
| Worthless | 0.808 | 0.808 | 0.808 | 0.808 | 0.808 | 0.808 |
| Depression | 0.864 | 0.864 | 0.864 | 0.864 | 0.864 | 0.864 |

*Outcome variable codes*
Lonely: RA1SA20G; Rel. Diff.: RA1SF1J
Worthless: RA1SA20F; Depression: RA1PDEPRE
*Predictor variable codes*
Age: RA1PRAGE\_ORD; Bill Diff.: RA1SH9; Purpose: RA1SF100
Self-Efficacy: RA1SF4F; Sleep: RA1SA57A; Sex: RA1PRSEX

Table 5 compares the four-predictor joint specification (`RA1PRAGE_ORD`, `RA1SH9`, `RA1SF100`, and `RA1SA57A`) against the corresponding single-predictor accuracy band. Joint modeling yields only a slight gain for relationship difficulty (0.310 vs. 0.291–0.304) and worthlessness (0.812 vs. 0.808), a negligible change for loneliness (0.654 vs. 0.653), and virtually identical accuracy for depression (0.863 vs. 0.864). These modest shifts underscore that rank association and hard top-1 class accuracy capture different facets of ordinal structure.

**Table 5:** Mean Stratified 5-Fold Cross-Validation Accuracy for Joint Four-Predictor Probit Models.

| **Outcome Variable** | **Joint CV**<br>**Accuracy** | **Single-Predictor**<br>**Range** |
|---|---|---|
| Felt lonely (RA1SA20G) | 0.654 | 0.653–0.653 |
| Relationship difficulty (RA1SF1J) | 0.310 | 0.291–0.304 |
| Felt worthless (RA1SA20F) | 0.812 | 0.808–0.808 |
| Depression (RA1PDEPRE) | 0.863 | 0.864–0.864 |

## 5. Discussion: Substantive Meaning and Methodological Trade-Offs

Taken together, the exploratory summaries, rank correlations, cumulative gradients, and probit evaluations depict midlife mental health as multi-faceted rather than reducible to a single determinant. Loneliness in particular has been prospectively linked to adverse mental health outcomes (13). Economic strain from monthly bills, sleep onset difficulty, and internal psychosocial resources such as purpose and self-efficacy each show clear rank alignment with emotional distress in MIDUS Project 1. Sleep disturbance has also been examined longitudinally alongside job strain (14). Yet individual survey items explain only a bounded share of classification variance, reflecting both measurement noise in single-item self-reports and broader life-course influences.

High cross-validated accuracy for depression (0.864) and worthlessness (0.808) reflects strong category concentration in the baseline low-severity tier (86.4% and 80.8%, respectively). Under hard arg max decision rules, predicting the dominant class already captures a large fraction of observations when tail categories are uncommon. Skewed category distributions of this kind are also frequent in biopharmaceutical outcome data (15). Conversely, relationship difficulty (`RA1SF1J`) spreads more evenly across intermediate agreement and disagreement levels, making exact category matching harder (0.291–0.310) even when monotonic rank gradients are visible.

Recoding choices illustrate a central trade-off in ordinal survey analysis between statistical stability and fine-grained resolution. Collapsing sparse disagreement tiers, merging upper frequency labels, trichotomizing the 0–7 depression score, and binning age into quartiles prevent empty or ultra-sparse cells during estimation. At the same time, merging categories blurs subtle distinctions near scale boundaries, and alternative cutpoints— such as clinically calibrated depression thresholds—could alter prevalence tiers and downstream estimates.

Choosing among rank correlation indices also requires careful attention to measurement scale and tie structure. Kendall’s τ_*b*_ handles tied ranks symmetrically for square ordinal tables, Stuart’s τ_*c*_ adjusts for rectangular table dimensions when category counts differ, and the rank-biserial coefficient summarizes rank separation across binary sex groups. All three coefficients quantify monotonic co-movement rather than causal effect, and the cross-sectional Project 1 design cannot establish temporal ordering among predictors and outcomes.

Several modeling assumptions warrant caution when interpreting the ordinal probit results. Entering ordered predictor codes as evenly spaced integers assumes equal latent steps between adjacent categories, while respondent sex enters as a nominal indicator rather than an ordered variable. Anxiety (`RA1PANXIE`) was left out of the post-recode outcomes because it remained numeric rather than binned, and evaluating models solely via hard classification accuracy ignores whether misclassifications land one step away or across multiple severity tiers. Explainability-oriented evaluation of this kind is similarly emphasized in NLP classification work (16).

Comparing single-predictor and joint specifications further highlights the distinction between out-of-sample hard accuracy and covariate adjustment. Adding four predictors simultaneously provides marginal gains in top-1 accuracy for relationship difficulty and worthlessness while leaving loneliness and depression nearly unchanged. Even when classification accuracy rises only slightly, multivariable ordinal probit models remain useful for examining adjusted associations across simultaneous stressors and resources.

## 6. Conclusion and Future Directions

This study presented an end-to-end ordinal analysis of MIDUS Project 1 survey items examining loneliness, relationship difficulty, worthlessness, and depression alongside demographic and psychosocial predictors. Starting from 3, 577 initial records and retaining 2, 423 complete cases, the workflow combined univariate profiling, theoretically guided category recoding, rank correlation analysis, cumulative exceedance curves, and cross-validated ordinal probit models.

Key empirical findings show substantial skewness toward low-symptom categories, consistent monotonic gradients linking financial strain and sleep trouble to higher outcome thresholds, and marked differences in classification accuracy across out-comes (0.864 for depression and 0.808 for worthlessness versus 0.291–0.310 for relationship difficulty). Joint four-predictor specifications offer only modest improvements in hard classification accuracy over single-predictor models, underscoring the limits of point-prediction accuracy on skewed ordinal scales.

Several avenues can advance this line of research in subsequent work. Incorporating survey weights would align sample estimates more closely with national population inference, while modeling anxiety (`RA1PANXIE`) directly on its numeric scale would preserve its full distributional range. Future analyses should also adopt ordinal-aware evaluation metrics such as quadratic weighted kappa, test proportional-odds and threshold assumptions formally, and leverage longitudinal MIDUS waves to evaluate within-person trajectories and strengthen causal inference.

## Data Availability

All data used in the present study are from the publicly available, de-identified MIDUS (Midlife in the United States) survey data. No new data were collected.

https://www.midus.wisc.edu/

